# Systematic Review and Meta-Analysis of the Associations Between Body Mass Index, Prostate Cancer, Advanced Prostate Cancer and Prostate Specific Antigen

**DOI:** 10.1101/19005421

**Authors:** Sean Harrison, Kate Tilling, Emma L. Turner, Richard M. Martin, Rosie Lennon, J. Athene Lane, Jenny L. Donovan, Freddie C. Hamdy, David E. Neal, J.L.H. Ruud Bosch, Hayley E. Jones

## Abstract

**Purpose:** The relationship between body-mass index (BMI) and prostate cancer remains unclear. However, there is an inverse association between BMI and prostate-specific antigen (PSA), used for prostate cancer screening. We conducted this review to estimate the associations between BMI and (1) prostate cancer, (2) advanced prostate cancer, and (3) PSA.

**Methods:** We searched PubMed and Embase for studies until 02 October 2017 and obtained individual participant data from four studies. In total, 78 studies were identified for the association between BMI and prostate cancer, 21 for BMI and advanced prostate cancer, and 35 for BMI and PSA. We performed random-effects meta-analysis of linear associations of log PSA and prostate cancer with BMI and, to examine potential non-linearity, of associations between categories of BMI and each outcome.

**Results:** In the meta-analyses with continuous BMI, a 5 kg/m^2^ increase in BMI was associated with a percentage change in PSA of −5.88% (95% CI −6.87% to −4.87%). Using BMI categories, compared to normal weight men the PSA levels of overweight men were 3.43% lower (95% CI −5.57% to −1.23%), and obese men were 12.9% lower (95% CI −15.2% to −10.7%). Prostate cancer and advanced prostate cancer analyses showed little or no evidence associations.

**Conclusion:** There is little or no evidence of an association between BMI and risk of prostate cancer or advanced prostate cancer, and strong evidence of an inverse and non-linear association between BMI and PSA. The association between BMI and prostate cancer is likely biased if missed diagnoses are not considered.

## 1. Background

Prostate cancer is the second commonest male cancer worldwide, (1) and the most commonly diagnosed cancer in men in the UK, with an estimated 47,151 diagnoses in 2015 (2). Generally, most prostate cancers are slow growing, but can metastasise to the bones, lungs and brain. Worldwide, there were an estimated 307,000 deaths from prostate cancer in 2012 (1), and in the UK, around 11,600 men died from prostate cancer in 2016 (2).

Body mass index (BMI) has been associated with many cancers (3), but its association with prostate cancer is unclear. Previous meta-analyses and reviews have suggested that BMI is not associated with prostate cancer (4,5), positively associated with prostate cancer (6,7) inversely associated with localized prostate cancer (8), and positively associated with advanced (8), aggressive (9), high-grade and fatal prostate cancers (4). These meta-analyses were either limited to cohort studies (5,7,8,10) or in need of updating (6,7). Additionally, no meta-analysis assessed potential non-linear associations between BMI and risk of prostate cancer or advanced prostate cancer. We therefore sought to perform an updated review of the literature, including more studies, and additionally examining non-linear associations.

BMI has also been inversely associated with prostate-specific antigen (PSA) (11), although no previous meta-analysis of this relationship exists. The presence of such an association could bias observed relationships between BMI and prostate cancer as PSA testing often plays a key role in diagnosis. More specifically, a negative association between BMI and PSA could lead to a spurious negative association or mask a positive association between BMI and localised prostate cancer, as obese men, with lower PSA values, would be less likely to be offered a biopsy as the result of a PSA test. A negative association between BMI and PSA could also induce a spurious positive association between BMI and advanced prostate cancer, as obese men may be diagnosed later, due to their lower PSA levels. In addition, if the association between BMI and prostate cancer (or advanced prostate cancer) is non-linear, then studies with different distributions of BMI will give rise to different estimates of the BMI-prostate cancer association.

We systematically reviewed the literature for all relevant studies and performed meta-analyses. We also examined these relationships using individual participant data (IPD) from prostate cancer studies. In analysing the IPD studies, we aimed to account for incomplete and PSA-dependent diagnosis by imputing prostate cancer status for all men who did not receive a biopsy, and in doing, avoid potential bias resulting from an association between BMI and PSA.

Our objectives were to: i) precisely quantify the (assumed linear) associations between BMI and prostate cancer, advanced prostate cancer and PSA; ii) update previous meta-analyses using all relevant evidence, including case-control studies; and iii) explore potential non-linearity in these associations. Our overall aim was to understand whether BMI is a risk factor for prostate cancer, and to identify whether failure to account for the role of PSA in many prostate cancer diagnoses is likely to lead to biased estimates of the association between BMI and prostate cancer.

## 2. Methods

### 2.1 Eligibility criteria

We performed a systematic review in which we included original articles published in peer reviewed journals that measured an association between BMI and total prostate cancer incidence and/or advanced prostate cancer; and studies that measured an association between BMI and PSA, including supplements and meeting abstracts; human randomised controlled trials (RCTs), case-control, cohort, cross-sectional, and non-randomised experimental studies. If the abstract did not specifically mention BMI but mentioned height or weight, we acquired the full text to determine if BMI was calculable from data included in the publication.

We excluded reviews, books, commentaries, letters, and animal and cell-line studies; studies examining pre-malignant disease if there was no mention of prostate cancer or PSA; studies where BMI was measured after diagnosis of prostate cancer, as this increases the likelihood of reverse causality; and studies that we considered to be at critical risk of bias (see **Section 2.4**).

We determined the effect estimate to be for advanced prostate cancer if the individual studies labelled the effect as “advanced” or “aggressive”, or if the effect was for locally advanced, extra-prostatic, nodular or metastatic prostate cancer. High-grade prostate cancer on its own was not considered equivalent to advanced prostate cancer and was not extracted.

### 2.2 Data Sources

We searched Medline and Embase databases up to 02 October 2017 for studies in humans associating BMI with either prostate cancer or PSA. The search query was as follows (each term as a text word search): (BMI or body-mass index or obese or obesity or body weight or body size or adiposity) AND (prostate cancer or prostate neoplasm or PSA or prostate-specific antigen) NOT psoriatic arthritis. Psoriatic arthritis was excluded as its initialism is also PSA. We also reviewed the reference lists of previous meta-analyses for further studies for inclusion (6,8,12). Duplicate studies were removed prior to download using the Ovid deduplication tool.

### 2.3 Data extraction

One author (SH) screened the titles and abstracts of all papers for inclusion and retrieved full texts for all studies that met the inclusion criteria. Full texts were also sought if no abstract was available or if the abstract did not include sufficient information to decide on inclusion. We also sought full texts for conference abstracts, if a corresponding full text was not found in the original search. If no full text could be found, and the abstract provided insufficient information for inclusion, the study was excluded. We excluded one published paper where we could not locate a full text (13).

One author (SH) screened all full texts for inclusion, and one of three independent reviewers (KT, ET, HJ) reviewed the first 60 full texts to check for consistency. We resolved any inconsistency with discussion to clarify screening criteria. A random subset of the remaining studies (30 full texts) were also reviewed by the independent reviewers to check for drift from inclusion/exclusion criteria.

Both SH and RL independently extracted all relevant data from included studies, with disagreements resolved by discussion. The first ten extractions were also performed by HEJ, KT and ELT to check for consistency.

We categorised prostate cancer studies as “before” if BMI was measured on average at least two years before diagnosis (prospective studies), and “same time” if BMI was measured on average less than two years before diagnosis. In general, “before” studies were cohort studies and “same time” studies were case-control studies. We considered the “before” studies to be at lower risk of reverse causation.

We extracted data that were (or could be transformed to) an odds ratio (OR) or hazard ratio (HR) quantifying the continuous association between BMI and total and advanced prostate cancer risk, and a regression coefficient for the association between BMI and log-PSA. Log-PSA was used as an outcome rather than PSA as we assumed a multiplicative association between BMI and PSA, which fits with the theory that haemodilution is responsible for any observed association (14). Studies reported associations in a variety of ways; a detailed list of the statistical conversions used to estimate the ORs, HRs and regression coefficients and their standard errors (SEs) is in **Supplementary appendix 1**.

We estimated linear associations, taking BMI as a continuous exposure variable, and assessing the possibility of non-linear associations by coding BMI as a categorical exposure. Specifically, we estimated linear associations between BMI and the log odds of prostate cancer or advanced prostate cancer, and between BMI and log transformed PSA. For simplicity, we refer to linear associations as “continuous” throughout. The following BMI categories were used: normal weight (BMI < 25 kg/m^2^), overweight (25 kg/m^2^ ≤ BMI < 30 kg/m^2^), and obese (BMI ≥ 30 kg/m^2^). We refer to these as “categorical” associations throughout.

When several papers reported on the same study, for continuous associations we prioritised papers that presented continuous effect estimates (e.g. HR or OR per 1 kg/m^2^ increase in BMI) over papers presenting categorical effect estimates (e.g. HR or OR for overweight and obese groups versus normal weight), and these were prioritised over mean differences. For categorical associations, we extracted estimates from papers presenting categorical associations only. If duplicate studies presented the same effect estimate types in multiple papers, the paper with the largest number of participants was used in the meta-analysis. If both adjusted (e.g. for potential confounders such as age, ethnicity, etc.) and unadjusted results were given in the same paper, the most-adjusted model was used in the meta-analysis.

If the data were insufficient to estimate a regression coefficient, OR or HR and SE, we extracted a P value, the number of participants and direction of association from the most relevant analysis for use in an albatross plot (15).

### 2.4 Risk of bias assessment

SH and RL assessed the risk of bias in each study independently using an assessment tool created for a previous meta-analysis (16), with disagreements resolved by discussion. This tool uses the categories of assessment from a draft of the ROBINS-I tool (17), and questions from the CASP case-control and cohort questionnaires (18,19), see **Supplementary appendix 2**.

We assessed risk of bias in six categories: confounding, selection of participants, missing data, outcome measurement, exposure measurement and results’ reporting. We assigned overall and category-specific risks of bias: either low, moderate, high, critical or unclear (if there was insufficient information to assign a risk). We based the overall risk of bias on a subjective combination of the category-specific risk of biases, looking at the maximum risk of bias that could have been introduced into the study by each category. The overall risk of bias was not low in any study, as all studies were observational and thus potentially subject to unmeasured confounding.

We determined that a study had a critical risk of bias if: i) age was not accounted for in either the design or analysis of the study and, for BMI-prostate cancer case control studies, if there was more than a 3-year difference in the mean or median ages of cases and controls, because age is strongly associated with BMI (20), prostate cancer risk (21), and PSA (21); or ii) if the design of the study was such that participation was conditional upon PSA levels, both for the association between BMI and PSA (as this would involve conditioning on the outcome) and the association between BMI and prostate cancer (as this would involve conditioning on a collider) (22).

Studies with a critical risk of bias were excluded prior to analysis and were not considered further.

In the studies found in the systematic review, it was generally unclear whether men considered as not having prostate cancer had received biopsies. Usually, the controls were “not known to have prostate cancer”, rather than “known not to have prostate cancer”. Therefore, screening could have introduced bias in the association between BMI and prostate cancer. Although we did not consider this a critical risk of bias, we sought to investigate and quantify this bias using large studies where biopsy status was known, and IPD available.

### 2.5 Individual participant data studies

Studies that offered prostate biopsies if the participants’ PSA were above threshold values (screening studies) were excluded from our systematic review for having a critical risk of bias. However, we noted that some of the largest potentially relevant studies for our research questions were screening studies, and that bias due to screening could potentially be accounted for using imputation of prostate cancer status if IPD were available. This would then allow these studies to be included in the meta-analyses.

We approached four prospective studies looking at prostate cancer to obtain IPD: Krimpen (23), Prostate Cancer Prevention Trial (PCPT) (24), Prostate, Lung, Colorectal, and Ovarian cancer screening trial (PLCO) (25) and Prostate Testing for cancer and treatment trial (ProtecT) (26). These studies were chosen because they were large studies of prostate cancer with known PSA screening protocols, or the biopsy status of all participants was known. Key to informing the imputation model was PCPT, which offered biopsies to all participants regardless of PSA level. This information allowed us to impute prostate cancer status for men with a PSA level below the threshold for biopsy in the other three studies. However, PCPT only included men with a PSA less than 3.0 ng/ml, biasing both the BMI-PSA and BMI-prostate cancer analyses, and as such was excluded from the meta-analyses.

For each IPD study, we requested data measured at baseline on BMI and PSA, as well as age, family history of prostate cancer and ethnicity. We also requested data on prostate cancer status (including tumour, node, metastases [TNM] and Gleason scores). For each man who was not biopsied, we imputed prostate cancer status by the end of the study in which he participated using multiple imputation. We included baseline age, BMI, log-PSA, family history of prostate cancer, and study as explanatory variables to impute prostate cancer status using logistic regression. BMI, log-PSA and family history of prostate cancer were also imputed if missing.

In each of the three included IPD studies, we estimated associations between BMI and (1) prostate cancer, (2) advanced prostate cancer and (3) PSA. We restricted the analyses to men with white ethnicity (due to low numbers of non-white men and therefore difficulties in imputation), and adjusted the analyses for age, family history of prostate cancer (for prostate cancer analyses), and prostate cancer status (for the PSA analyses). Full details of the IPD studies, the imputation method and statistical analyses are available in Supplementary Appendix 3.

### 2.6 Combining data

#### Meta-analysis

We combined estimates from studies identified through the systematic review and the IPD studies using random-effects and fixed effect meta-analyses. We performed separate meta-analyses of continuous and categorical associations for each outcome (prostate cancer, advanced prostate cancer and PSA). All meta-analysis results are presented in forest plots.

Studies presenting HRs and ORs were analysed and presented separately. For studies presenting ORs, “same time” and “before” studies were meta-analysed in subgroups, and labelled as such in forest plots. Studies presenting HRs were all classed as “before” studies, and labelled simply “HR”. The results are presented as the HR or OR for prostate cancer or advanced prostate cancer and percentage change in PSA for a 5 kg/m^2^ increase in BMI. Heterogeneity was tested for and quantified using the Cochran’s Q and I^2^ statistics (27,28).

In meta-analyses of categorical associations, studies from the systematic review were included if they presented ORs or HRs for overweight and/or obese men relative to normal weight men (for the outcomes of prostate cancer and advanced prostate cancer) or means and SDs of PSA or log-PSA for each of these BMI categories (for the outcome of PSA). ORs and HRs that were presented for other categories of BMI were not used (such as morbidly obese, BMI ≥ 35 kg/m^2^), though we combined the mean and SD of PSA for different categories with neighbouring categories when sufficient information was available.

#### Meta-regression

Meta-regression (29) was used to determine if the effect estimates from individual studies included in the meta-analyses varied by study-level factors. For all meta-regressions, we considered ethnicity (non-white versus white in each study, defined as >80% white participants or from a country with a majority white population), mid-year of recruitment, mean BMI in the study, and the overall risk of bias (high versus medium). For the associations between BMI and prostate cancer and advanced prostate cancer, we also considered the mean age at diagnosis, and study mean time between BMI measurement and diagnosis.

#### Funnel plots

Funnel plots (30) were drawn to assess for small study effects in each analysis (31).

#### Albatross plots

As not all studies reported enough information to be included in the meta-analyses, we also present albatross plots containing results from studies with and without sufficient information to be included in the meta-analyses (32). These are plots of the P value of an association against the number of participants and can be used to assess heterogeneity between studies and assess the rough magnitude of an association using limited information. By indicating which studies had insufficient data to contribute to meta-analysis on the albatross plots, we determined whether inclusion of the remaining studies would have altered the overall interpretation of the evidence.

## 3. Results

In total, **9**,**127** papers were found that had keywords for BMI and prostate cancer or PSA. After title and abstract screening, **725** papers remained (see **Figure 1**, PRISMA flow diagram). After full text screening, risk of bias assessment, and removal of papers reporting the same studies, **78** studies examined the association between BMI and prostate cancer (**67** with data for meta-analysis), **21** studies examined the association between BMI and advanced prostate cancer (**18** with data for meta-analysis), and **35** studies examined the association between BMI and PSA (**20** with data for meta-analysis, **one** of which only had data for categorical associations).

**Figure 1.**
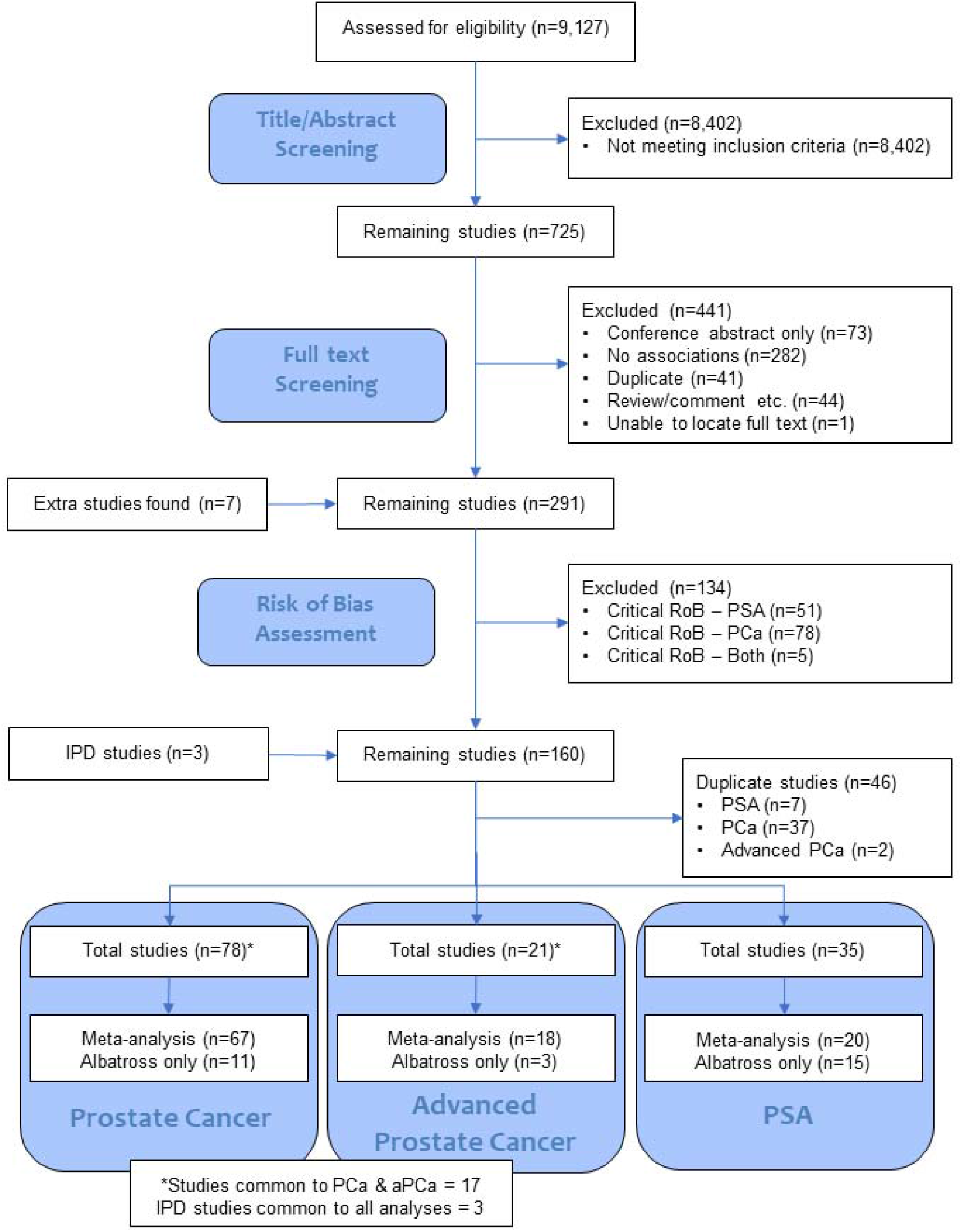
PRISMA flow diagram showing the number of studies in each stage of the systematic review.

A summary of all results is given in **Box 1**.

### 3.1 BMI and Prostate Cancer

#### Continuous BMI

Of the 78 studies examining the association between BMI and prostate cancer (23,25,26,33–107), 11 (14%) could not be included in the meta-analysis due to insufficient data but were included in the albatross plot (97–107). All studies are detailed in **Supplementary Table 1**, with the results of the risk of bias assessment in **Supplementary Table 2**.

In total, 9,513,326 men from 67 studies were included in the HR and OR meta-analyses, (9,351,795 in 30 HR studies, 161,531,383 in 37 OR studies); of these, 201,311 (2.1%) men had prostate cancer (157,990 cases [1.7%] in HR studies, 41,863 [25.9%] in OR studies). The random-effects meta-analyses (**Figures 2 and 3**) estimated the average HR and OR for prostate cancer for a 5 kg/m^2^ increase in BMI to be 1.01 (95% CI 0.99 to 1.04, P = 0.29) and 0.99 (95% CI 0.96 to 1.02, P = 0.58) respectively. There was strong evidence for heterogeneity in effect estimates across studies for the studies reporting an HR (P < 0.001, I^2^ = 79.9%), and studies reporting an OR (P < 0.001, I^2^ = 66.1%). Pooled estimates from fixed effect meta-analyses were essentially the same.

**Figure 2.**
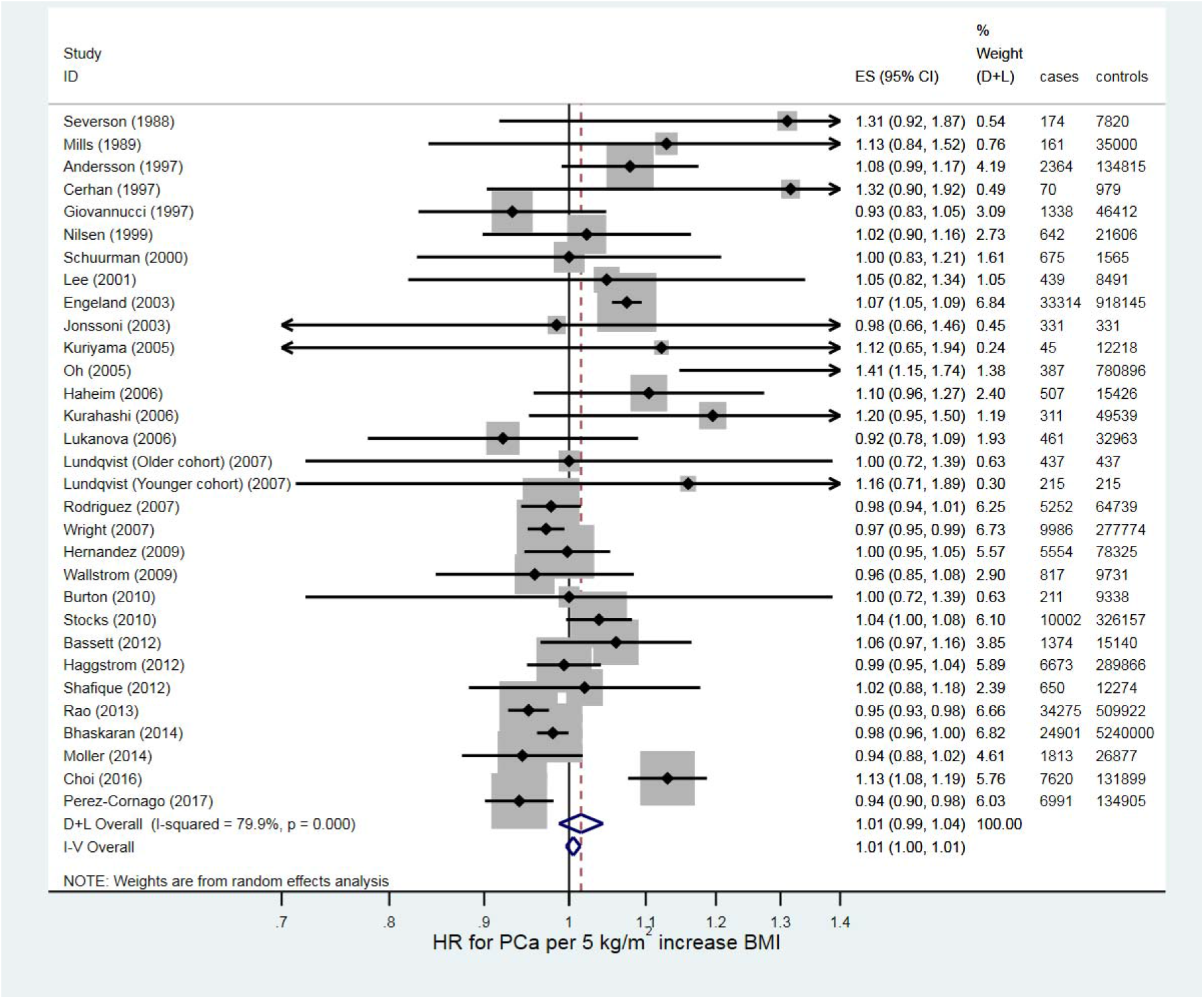
Forest plot for the association between BMI and prostate cancer (hazard ratios)

**Figure 3.**
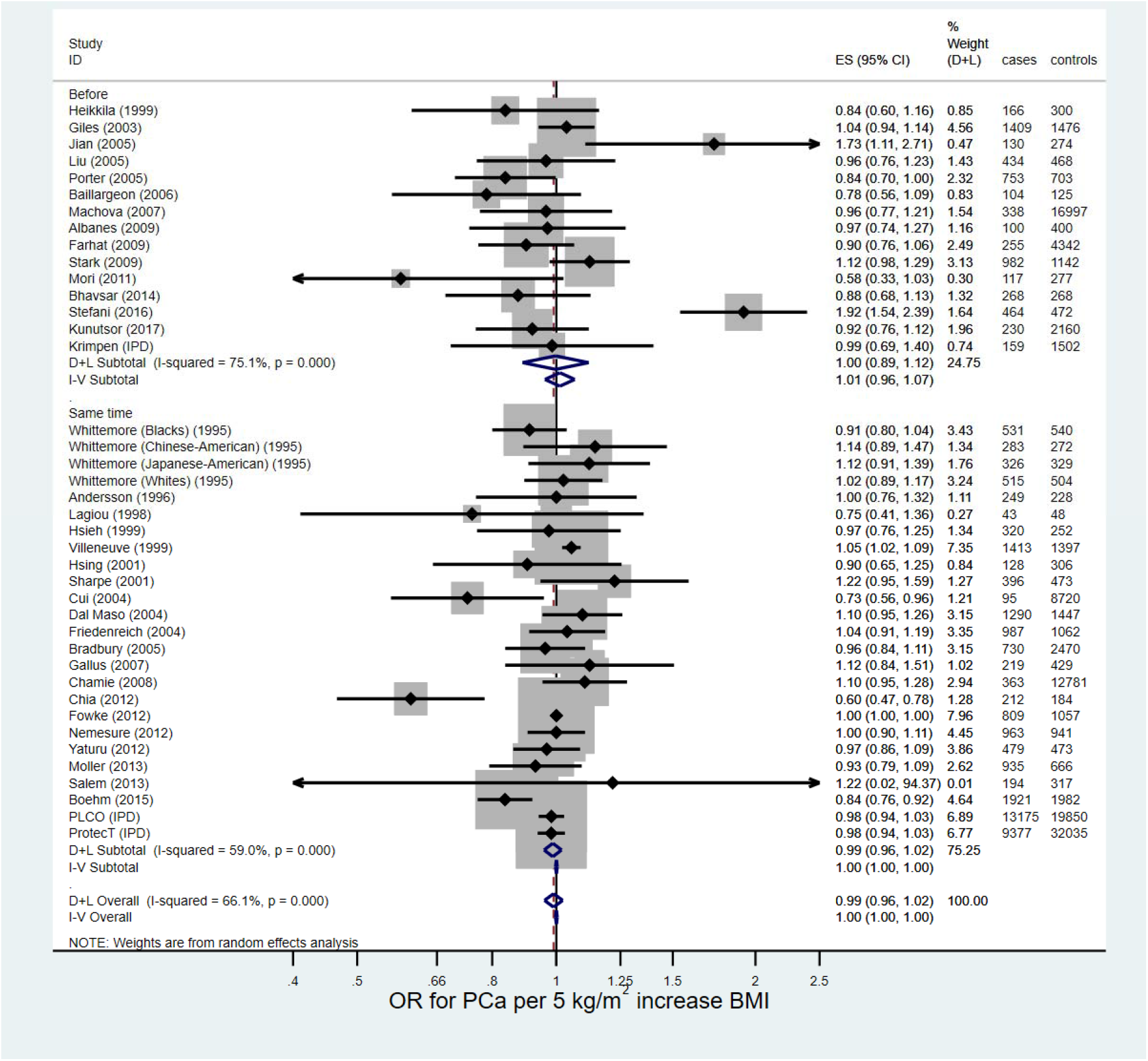
Forest plot for the association between BMI and prostate cancer (odds ratios)

From a meta-analysis including only IPD studies, the estimated average OR for prostate cancer for a 5 kg/m^2^ increase in BMI was 0.98 (95% CI 0.95 to 1.01) (**Supplementary Appendix 3.6**). Analysed without imputation (complete case analysis), the estimated OR was only 0.94 (95% CI 0.91 to 0.97).

There was limited evidence of (positive) small study effects on the funnel plot for HRs, but not ORs (**Supplementary Figures 1 and 2**). The albatross plot (Supplementary Figure 3) showed that the eleven studies without sufficient information for meta-analysis were spread evenly across both positive and negative effect sizes, consistent with the null result seen in the meta-analysis.

Meta-regression (**Supplementary Table 3**) on study-level variables did not explain any of the heterogeneity.

#### Categorical BMI

Thirteen of the studies included in the continuous meta-analyses above presented HRs or ORs for overweight and/or obese men versus normal weight men (23,25,26,42,49,50,57,66–69,75,108). Only ten studies presented HRs or ORs for overweight men, whereas all thirteen presented HRs or ORs for obese men versus normal weight men. In total, there were 252,771 participants and 32,277 men with prostate cancer included in this meta-analysis; two studies (50,108) did not report how many men were in each BMI subgroup and were not included in these totals.

**Supplementary Table 4** shows the mean BMI, total number of men and number of men with prostate cancer in each category of BMI, and **Supplementary Table 5** shows the HRs and ORs for prostate cancer for each study for overweight and obese versus normal weight men. Forest plots are presented in **Supplementary Figures 4 and 5**. For the random-effects meta-analysis, the average HR for prostate cancer between overweight and normal weight men was estimated to be 1.02 (95% CI 0.98 to 1.05, P = 0.35) with no evidence of heterogeneity (I^2^ = 0.0%, P = 0.66), and the average OR was estimated to be 0.99 (95% CI 0.91 to 1.08, P = 0.89, combined across ORs for BMI measured before and at the same time as prostate cancer diagnosis) with little evidence of heterogeneity (I^2^ = 34.7%, P = 0.18). The average HR for prostate cancer between obese and normal weight men was estimated to be 0.97 (95% CI 0.93 to 1.01, P = 0.16), with no evidence of heterogeneity (I^2^ = 0.0%, P = 0.80), and the average OR was estimated to be 0.90 (95% CI 0.81 to 1.00, P = 0.05, combined across ORs), with some evidence of heterogeneity (I^2^ = 41.3%, P = 0.10). Fixed-effect models gave very similar results.

**Figure 4.**
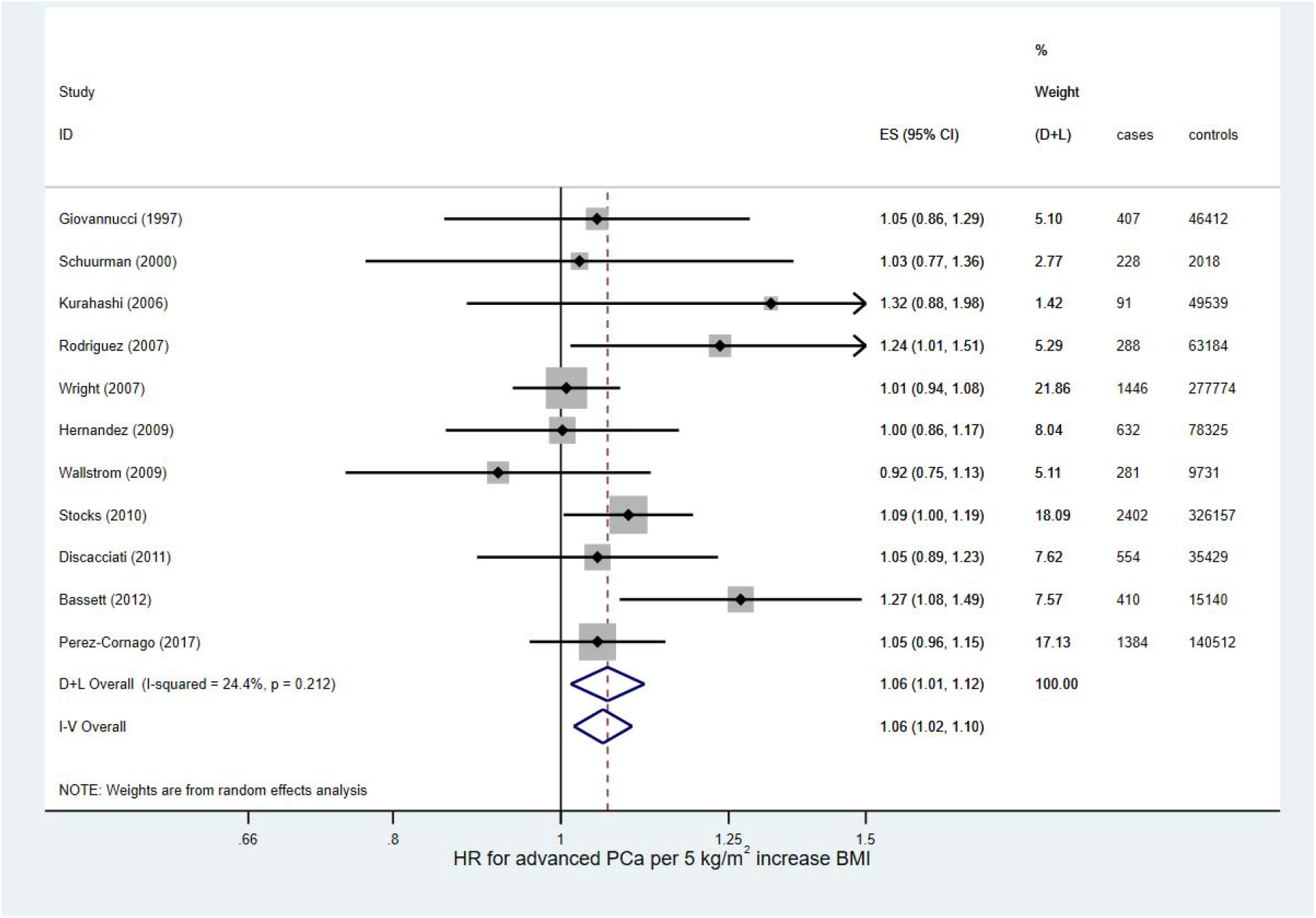
Forest plot for the association between BMI and advanced prostate cancer (hazard ratios)

**Figure 5.**
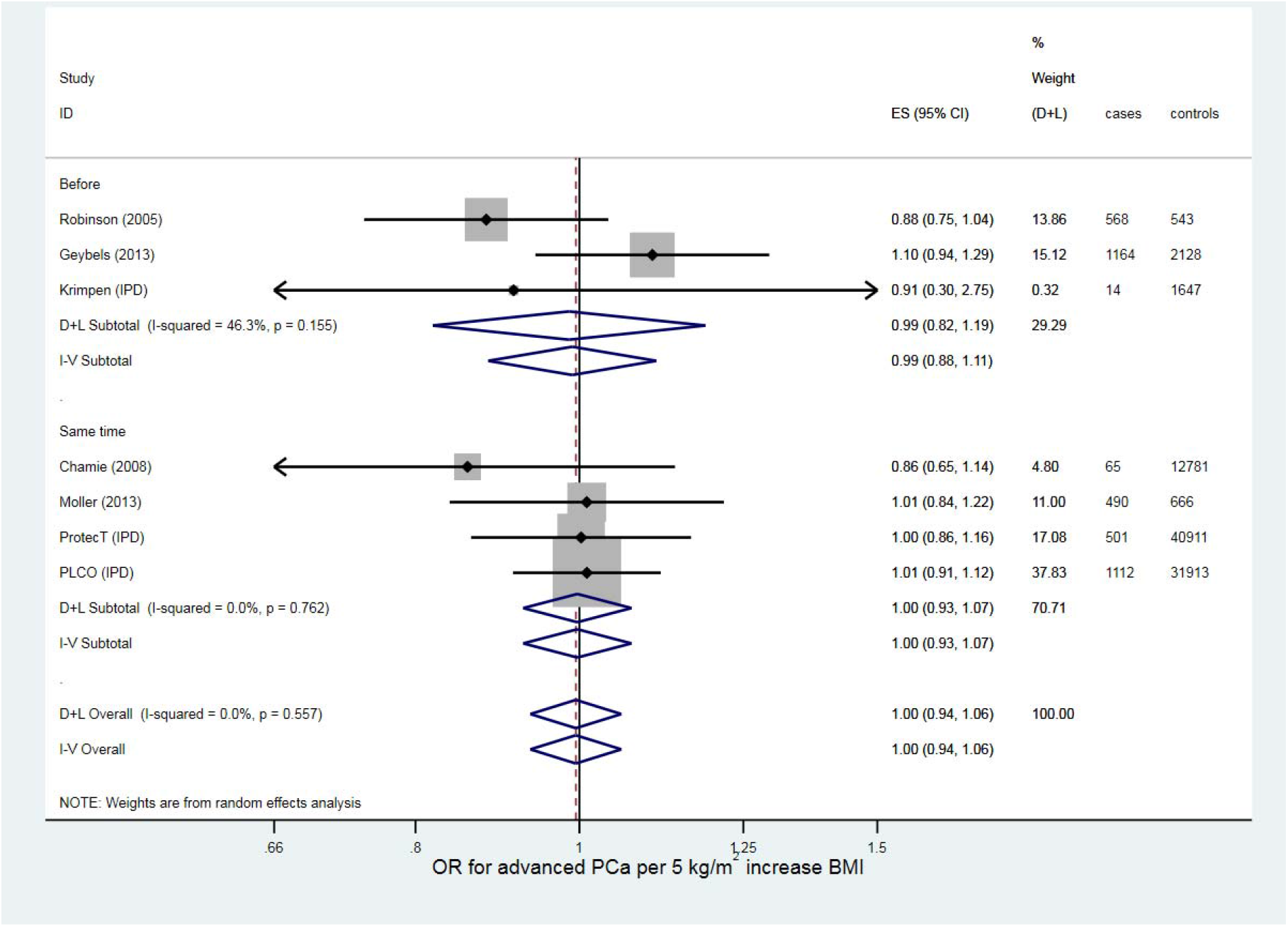
Forest plot for the association between BMI and advanced prostate cancer (odds ratios)

The heterogeneity in the average OR for prostate cancer between obese and normal weight men may have been due to differences between IPD and non-IPD studies. There was no evidence of heterogeneity for either IPD (OR = 0.97, 95% CI 0.91 to 1.04, P = 0.46) or non-IPD (OR = 0.77, 95% CI 0.67 to 0.89, P < 0.001) studies when considered separately (I^2^ = 0.0% for both, P = 0.93 and P = 0.54 respectively).

### 3.2 BMI and Advanced Prostate Cancer

#### Continuous BMI

Of the 21 studies examining the association between BMI and advanced prostate cancer (23,25,26,37,39,46,49–52,54,55,62,90,95,100,101,104,109–111), 3 studies (14%) could not be included in the meta-analysis due to insufficient data but were included in an albatross plot (100,101,104). The studies examining the association between BMI and advanced prostate cancer are detailed in **Supplementary Table 6**, with the results of the risk of bias assessment in **Supplementary Table 7**.

In total, 1,146,847 men were included from 18 studies (1,052,344 in 11 HR studies, 94,503 in seven OR studies); of these, 12,037 (1.0%) men had advanced prostate cancer (8,123 [0.8%] in HR studies, 3,914 [4.1%] in OR studies). The random-effects meta-analyses (**Figures 4 and 5**) estimated the average HR and OR for advanced prostate cancer for a 5 kg/m^2^ increase in BMI to be 1.06 (95% CI 1.01 to 1.12, P = 0.013) and 1.00 (95% CI 0.94 to 1.06, P = 0.88) respectively. There was little evidence for heterogeneity in effect estimates across studies reporting an HR (I^2^ = 24.4%, P = 0.21), and no evidence for heterogeneity in effect estimates across studies reporting an OR (I^2^ = 0.0%, P = 0.56). The fixed effect analysis showed essentially the same results.

When IPD studies were analysed separately, the estimated average OR for advanced prostate cancer for a 5 kg/m^2^ increase in BMI was 1.00 (95% CI 0.92 to 1.09), **Supplementary Appendix 3.6**. The effect estimate when analysed without imputation (complete case analysis) was slightly lower, with an estimated average OR of 0.98 (95% CI 0.89 to 1.08).

The funnel plots (**Supplementary Figures 6 and 7**) did not show evidence of any small study effects. The albatross plot (**Supplementary Figure 8**) showed that the three studies without sufficient information for meta-analysis all estimated a positive association between BMI and advanced prostate cancer risk. One small study of 1,474 men, Putnam (2000) (101), estimated an inconsistently strong effect. Because this study was so small, it does not change our interpretation of the meta-analyses.

**Figure 6.**
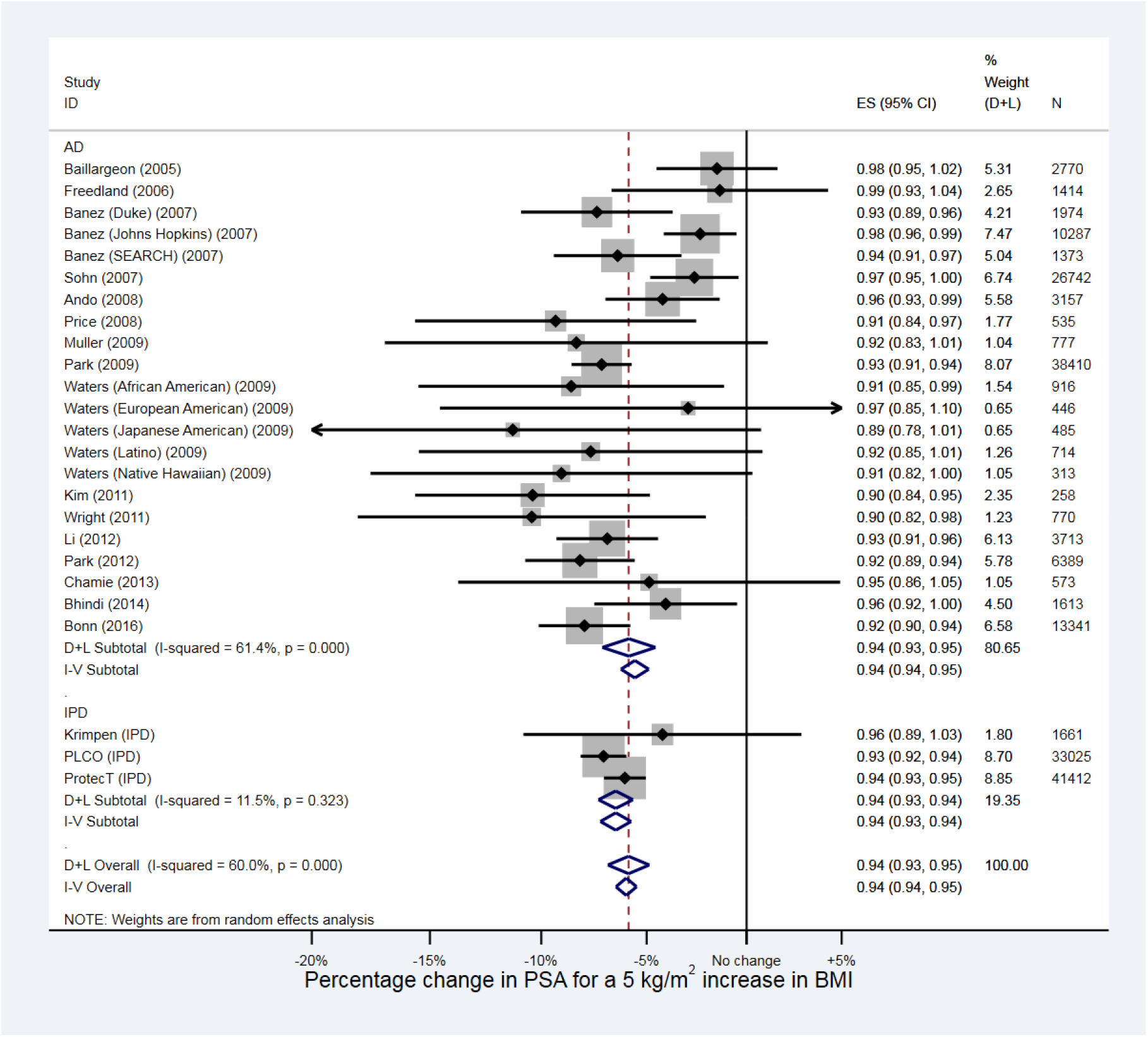
Forest plot for the association between BMI and PSA. AD = aggregate data, IPD = individual participant data.

Meta-regression (Supplementary Table 8) did not show evidence of any variation in results due to study-level variables.

#### Categorical BMI

Six of the studies included in the continuous meta-analysis presented HRs or ORs for overweight and/or obese men versus normal weight men (23,25,26,49–51). Only five studies presented results for overweight versus normal weight men, whereas all six presented results for obese versus normal weight men. In total, there were 169,530 participants included in this analysis, and 2,381 men had advanced prostate cancer (1.4%) (one study (50) did not report how many men were in each BMI subgroup and was not included in these totals).

**Supplementary Table 9** shows the mean BMI, total number of men and number of men with advanced prostate cancer in each category of BMI, and **Supplementary Table 10** shows the HRs and ORs for advanced prostate cancer, for each study for overweight and obese versus normal weight men. Forest plots are presented in **Supplementary Figures 9 and 10**. For the random-effects meta-analysis, the average HR for advanced prostate cancer between overweight and normal weight men was estimated to be 1.04 (95% CI 0.94 to 1.15, P = 0.44), with no evidence of heterogeneity (I^2^ = 0.0%, P = 0.74), and the average OR was estimated to be 1.05 (95% CI 0.89 to 1.25, P = 0.54), with no evidence of heterogeneity (I^2^ = 0.0%, P = 0.78). The average HR for advanced prostate cancer between obese and normal weight men was estimated to be 1.15 (95% CI 0.92 to 1.44, P = 0.22), with evidence of heterogeneity (I^2^ = 53.7%, P = 0.02), and the average OR was estimated to be 0.99 (95% CI 0.81 to 1.21, P = 0.92), with no evidence of heterogeneity (I^2^ = 0.0%, P = 0.68). Fixed-effect models gave very similar results.

### 3.2 BMI and PSA

#### Continuous BMI

Of the 34 studies providing information on the association between BMI (as a continuous variable) and PSA (23,25,26,112–142), 15 studies (42%) could not be included in the meta-analysis due to insufficient data but were included in an albatross plot (128–142). All included studies are detailed in **Supplementary Table 11**, with the results of the risk of bias assessment in **Supplementary Table 12**.

In total, 264,970 men from 19 studies were included in the meta-analysis. The random-effects meta-analysis (Figure 6) estimated the average percentage change in PSA for a 5 kg/m^2^ increase in BMI to be −5.88% (95% CI −6.87% to −4.87%, P < 0.001). There was strong evidence for heterogeneity in effect estimates across studies (I^2^ = 60.0%, P <0.001). The fixed-effect analysis showed essentially the same result with narrower confidence intervals (percentage change in PSA = −5.99%, 95% CI - 6.48% to −5.49%, P < 0.001).

The funnel plot (Supplementary Figure 11) showed little evidence of small-study effects. The albatross plot (Supplementary Figure 12) showed that the excluded studies were broadly consistent with the meta-analysis effect size.

Meta-regression (Supplementary Table 13) did not explain any of the observed heterogeneity.

#### Categorical BMI

Sixteen of the studies included in the continuous meta-analysis presented PSA or log-PSA levels for overweight and/or obese men and normal weight men (23,25,26,112–119,121,122,124,126,143), and one further study presented only categorical results (144). Overall, there were 17 studies and 218,700 participants included in this analysis.

**Supplementary Table 14** displays the average log-PSA in each BMI subgroup for all 17 included studies, and **Supplementary Table 15** displays the percentage MD in PSA for all comparisons. Forest plots are presented in **Supplementary Figures 13 and 14**. For the random-effects meta-analysis, the average percentage change in PSA between overweight and normal weight men was estimated to be −3.43% (95% CI −5.57% to −1.23%, P = 0.002), with strong evidence of heterogeneity across studies (I^2^ = 80.9%, P < 0.001), and the average percentage change in PSA between obese and normal weight men was estimated to be −12.9% (95% CI −15.2% to −10.7%, P < 0.001), with strong evidence of heterogeneity across studies (I^2^ = 69.5%, P < 0.001). The pooled estimates from fixed-effect meta-analyses were slightly lower for the change in PSA between overweight and normal weight men (percentage change = −2.56%, 95% CI −3.34% to −1.78%, P < 0.001), but similar for the change in PSA between obese and normal weight men (percentage change = −12.1%, 95% CI −13.2% to −11.1%, P < 0.001).

The difference in log-PSA between the obese and normal groups (−0.139) was almost four times the difference between the overweight and normal weight groups (−0.035). The weighted mean BMI across all studies was 22.2 kg/m^2^ for the normal BMI category, 26.5 kg/m^2^ for the overweight category, and 31.3 kg/m^2^ for the obese category. We therefore consider this evidence that there is a non-linear association between BMI and log-PSA.

## 4. Discussion

### Overall Prostate Cancer

There was no compelling evidence to suggest there is a linear association between BMI and prostate cancer risk, nor an association between being overweight and prostate cancer risk, and little evidence for an association between being obese and prostate cancer risk. However, there is likely a reduced risk of being diagnosed with prostate cancer in overweight/obese men due to the role of PSA screening or testing in many prostate cancer diagnoses. This is reflected in our analyses of the IPD studies: the complete case analysis in which we ignored the problem of incomplete diagnosis (not all men being biopsied) suggested a negative association between BMI and prostate cancer. This association was attenuated to the null after imputation of missing prostate cancer status in non-biopsied men. This finding is consistent with our hypothesis regarding the expected direction of bias due to the negative association of BMI with PSA.

Obese men with prostate cancer may also have a higher risk of missed diagnoses due to having larger prostates (145), which are associated with a lower likelihood of detecting prostate cancer at biopsy (146,147). Bias from PSA testing will be highest in populations with a high level of PSA screening. In other populations, obesity may affect the chance of receiving a PSA test, and therefore receiving a prostate cancer diagnosis, for example if obese men access primary care more.

Overall, our results are consistent with previous meta-analyses. A random-effects dose-response meta-analysis of prospective studies was conducted by Markozannes et al. (2016) (4) using data from the World Cancer Research Fund (WCRF) as part of the continuous update project (148). Markozannes included 39 studies with 3,798,746 participants and 88,632 men with prostate cancer (2.3%) for the association between BMI and prostate cancer (excluding studies on mortality), including many of the same studies we included in our meta-analysis. The pooled risk ratio (RR) for prostate cancer for a 5 kg/m^2^ increase in BMI was 1.00 (95% CI 0.97 to 1.03), consistent with our results. In addition, an umbrella review of systematic reviews and meta-analysis by Kyrgiou et al. (2017) (3) concluded that there was no strong evidence for an association between BMI and prostate cancer risk, with a summary OR for prostate cancer for a 5 kg/m^2^ increase in BMI of 1.03 (95% CI 0.99 to 1.06).

### Advanced Prostate Cancer

There was some evidence to suggest a linear association between BMI and the risk of advanced prostate cancer, but only among studies reporting an HR (HR = 1.06, 95% CI 1.01 to 1.12, P = 0.013). This association was not seen in studies reporting an OR, where most of the power came from the imputed IPD studies, indicating the possibility of bias in the HR estimate due to PSA screening. Additionally, there may be collider bias (22) from conditioning on prostate cancer, since any unmeasured confounders associated with both prostate cancer and advanced prostate cancer would induce an association between BMI and advanced prostate cancer. However, since the overall association between BMI and prostate cancer appears to be negligible, this is not a primary concern.

Markozannes conducted a meta-analysis of prospective studies of BMI and combined advanced, high-grade and fatal prostate cancer using WCRF data, which included 23 studies with 1,676,220 participants and 11,204 men with advanced/high-grade/fatal prostate cancer (0.67%) (4). The RR for advanced/high-grade/fatal prostate cancer for a 5 kg/m^2^ increase in BMI was 1.08 (95% CI 1.04 to 1.12). The effect estimate may be increased in the WCRF analysis by the inclusion of high-grade and/or fatal prostate cancers or exclusion of case-control studies. Kyrgiou et al. (2017) (3) concluded that there was weak evidence for a positive association between increasing BMI and advanced prostate cancer risk, with a RR for advanced prostate cancer for a 5 kg/m^2^ increase in BMI of 1.09 (95% CI 1.02 to 1.16), although our meta-analysis included more up-to-date studies with a stricter inclusion criteria.

### PSA

There was strong evidence of an inverse association between BMI and PSA, which we found to be likely non-linear, decreasing more quickly between overweight and obese than normal weight and overweight. On average, obese men have an estimated 12.9% lower PSA than a normal weight man, and overweight men 3.4% lower PSA. We could only find one previous review of the association between BMI and PSA, which did not include a meta-analysis or estimate effect size (158). Their conclusion was that many studies reported an inverse association between BMI and PSA, in agreement with our findings.

It could thus be beneficial to account for BMI when interpreting the results of a PSA test. One suggestion based on these results is to increase an overweight man’s PSA by 3.5% (multiply by 1.035) before comparing to a threshold, and an obese man’s PSA by 13% (11,149). As an example of the impact of doing so, 23% of men in ProtecT were obese, and 1.9% of these men had an observed PSA of less than 3.0 ng/ml, but a ‘corrected’ PSA above a 3.0 ng/ml threshold for biopsy when adjusted for the effect of BMI on PSA.

### Strengths and Limitations

We synthesised data from many studies, including participants from many different populations at different time points, improving generalisability. The total number of participants included in analyses was also very large, and as such all pooled effect estimates were precise. By including studies where BMI was measured before, and those where it was measured at the same time as prostate cancer detection, we could compare different study types: there was little difference between these two study types for all outcomes in the continuous analyses, suggesting the findings are robust to reverse causation of BMI change by PCa diagnosis. By including IPD studies and imputing prostate cancer status in men who were not biopsied, we were able to show and account for bias in the association between BMI and prostate cancer from PSA testing.

A further strength of this study was the inclusion of studies where only a P value and number of participants could be extracted, using albatross plots.

However, there are limitations. Many of the studies included in the meta-analysis compared men with a diagnosis of prostate cancer versus men without a diagnosis of prostate cancer. In the screening studies, most men were not biopsied. Assuming that none of these men had prostate cancer would be a strong assumption and likely lead to bias. We addressed this problem by treating prostate cancer status as missing in these men and using multiple imputation. We performed checks on the validity of our imputation model, but we note the limitation that our results may have been sensitive to the choice of this model. In the meta-analysis of all studies, we limited bias due to testing for prostate cancer with PSA by excluding studies that exclusively screened for prostate cancer (and thus would have the greatest bias), but as PSA screening is used in general practice the bias could not be entirely removed. The proportion of prostate cancers detected by testing with PSA likely varied in each study, potentially accounting for some of the heterogeneity in studies examining the association between BMI and prostate cancer and advanced prostate cancer. Indeed, all the heterogeneity between OR results for prostate cancer between obese and normal weight men was due to differences between the imputed IPD studies versus the non-IPD studies.

Overall, there were large amounts of heterogeneity between non-IPD studies in the continuous analyses of BMI and prostate cancer, and advanced prostate cancer. This could be due to heterogeneity across populations, methods of diagnosing prostate cancer, or differential adjustment for confounders in each study-specific analysis. Equally, because the studies may not have used the same definition of advanced prostate cancer, and because advanced prostate cancers could be locally advanced prostate cancer, nodes or metastatic cancer, these studies may be relatively heterogeneous. This may have attenuated any association between BMI and advanced prostate cancer.

There was also evidence of heterogeneity between studies examining the associations between BMI and PSA. As with the prostate cancer studies, the PSA studies adjusted for different confounders, therefore residual confounding may have increased heterogeneity. It is also possible the association between BMI and PSA varies by population, though our meta-regressions did not find any explanatory factors.

There was at least a moderate risk of bias for all studies, as all studies were observational and therefore could have been biased by unobserved confounding. We attempted to limit effects of bias by identifying key confounders and only including studies without a critical risk of bias. There was also no evidence from the meta-regression that the studies with a medium risk of bias had systematically different effect estimates than those with a high risk of bias.

In the categorical analyses, it was only possible to combine studies presenting results for specific categories of BMI. As such, relatively few studies were included; a superior approach would be to gather IPD from all eligible studies and to determine the precise form of any non-linear associations, which would also allow more accurate corrections to men’s PSA levels.

## 5. Conclusion

There was no evidence of an association between BMI and prostate cancer risk, and little evidence for an association with advanced prostate cancer risk. There was, however, strong evidence for an inverse non-linear association between BMI and PSA. There was evidence from IPD studies to suggest this could bias the association between BMI and prostate cancer in screening studies. Studies in populations where PSA testing is involved in diagnosis of prostate cancer should determine whether an exposure could be associated with PSA, and thus whether the observed association with prostate cancer could be biased.

## Data Availability

All summary data used is available on Github, linked in the paper

https://github.com/sean-harrison-bristol/BMI-PCa-PSA-meta-analysis

## Acknowledgments

ProtecT Support: The ProtecT trial is funded by the UK National Institute for Health Research (NIHR) Health Technology Assessment Programme (projects 96/20/06, 96/20/99, ISRCTN20141297) with the University of Oxford (Oxford, UK) as sponsor. The views and opinions expressed herein are our own and do not necessarily reflect those of the Department of Health. We acknowledge the tremendous contribution of all the ProtecT study participants, investigators, researchers, data monitoring committee, and trial steering committee. We acknowledge the support from the Oxford NIHR Biomedical Research Centre through the Surgical Innovation and Evaluation Theme and the Surgical Interventional Trials Unit, and Cancer Research UK through the Oxford Cancer Research Centre. This work was supported by Cancer Research UK project Grants C11043/A4286, C18281/A8145, C18281/A11326 and C18281/A15064 and a programme grant (the CRUK Integrative Cancer Epidemiology Programme, ICEP: C18281/A19169). The authors would like to acknowledge the support of the National Cancer Research Institute (NCRI) formed by the Department of Health, the Medical Research Council (MRC) and Cancer Research UK. The NCRI provided funding through ProMPT (Prostate Mechanisms of Progression and Treatment), and this support is gratefully acknowledged. The ProtecT funding source had no role in the design, conduct of the study, collection, management, analysis and interpretation or preparation, review, or approval of the article.

PCLO Support: The authors thank the Nation Cancer Institute for access to NCI’s data collected by the Prostate, Lung, Colorectal and Ovarian Cancer Screening Trial. The statements contained herein are solely those of the authors and do not represent or imply concurrence or endorsement by NCI.

PCPT Support: Research reported in this publication was supported in part by the National Cancer Institute of the National Institutes of Health under Award Numbers UM1CA182883 and U10CA37429. The content is solely the responsibility of the authors and does not necessarily represent the official views of the National Institutes of Health.

---

**Box 1 Summary of results**

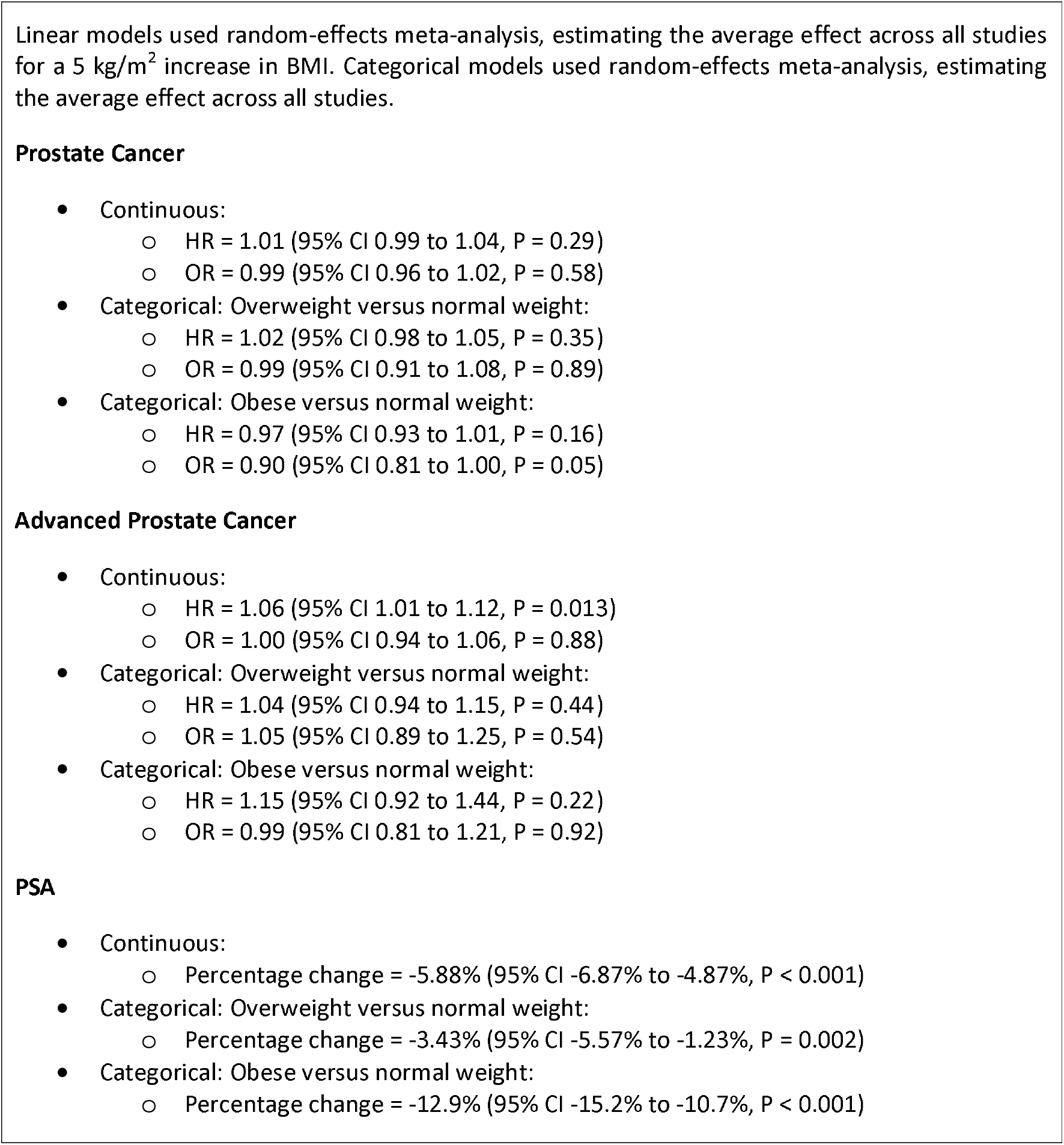

## Supplementary Figure Legends

Figure S1 Funnel plot for the association between BMI and prostate cancer (hazard ratios)

Figure S2 Funnel plot for the association between BMI and prostate cancer (odds ratios)

Figure S3 Albatross plot for the association between BMI and prostate cancer

Figure S4 Forest plot of the HRs and ORs for prostate cancer for overweight versus normal weight BMI categories, n1 = number of normal weight participants, n2 = number of overweight participants, blanks indicate missing data

Figure S5 Forest plot of the HRs and ORs for prostate cancer for obese versus normal BMI categories, n1 = number of normal weight participants, n3 = number of obese participants, blanks indicate missing data

Figure S6 Funnel plot for the association between BMI and advanced prostate cancer (hazard ratios) Figure S7 Funnel plot for the association between BMI and advanced prostate cancer (odds ratios) Figure S8 Albatross plot for the association between BMI and advanced prostate cancer

Figure S9 Forest plot of the HRs and ORs for advanced prostate cancer for overweight versus normal weight BMI categories, n1 = number of normal weight participants, n2 = number of overweight participants, blanks indicate missing data. ORs (before and same time) were combined as the Krimpen study was the only study measuring BMI before the outcome and presenting an OR

Figure S10 Forest plot of the HRs and ORs for advanced prostate cancer for obese versus normal weight BMI categories, n1 = number of normal weight participants, n3 = number of obese participants, blanks indicate missing data. ORs (before and same time) were combined as the Krimpen study was the only study measuring BMI before the outcome and presenting an OR

Figure S11 Funnel plot for the association between BMI and PSA

Figure S12 Albatross plot for the association between BMI and log-PSA. AD = aggregate data, IPD = individual participant data

Figure S13 Forest plot of the percentage change in PSA between overweight and normal BMI categories, n1 = number of normal weight participants, n2 = number of overweight participants, AD = aggregate data, IPD = individual participant data

Figure S14 Forest plot of the percentage change in PSA between obese and normal BMI categories, n1 = number of normal weight participants, n3 = number of obese participants, AD = aggregate data, IPD = individual participant data

